# Identifying patients at risk for myasthenic crisis with hemogram and inflammation-related laboratory parameters – a pilot study

**DOI:** 10.1101/2023.09.19.23295421

**Authors:** Anne Mehnert, Sivan Bershan, Jil Kollmus-Heege, Lea Gerischer, Meret Luise Herdick, Sarah Hoffmann, Sophie Lehnerer, Franziska Scheibe, Frauke Stascheit, Maike Stein, Alastair M. Buchan, Andreas Meisel, Annette Aigner, Philipp Mergenthaler

**Author notes:** Correspondence: Philipp Mergenthaler, Charité – Universitätsmedizin Berlin, Center for Stroke Research Berlin, Charitéplatz 1, 10117 Berlin, Germany. equal contribution (Annette Aigner, Philipp Mergenthaler).

## Abstract

**Background:** Myasthenia gravis (MG) is a rare autoimmune disease characterized by fatigable weakness of the voluntary muscles and can exacerbate to life-threatening myasthenic crisis (MC), requiring intensive care treatment. Routine laboratory parameters are a cost-effective and widely available method for estimating clinical outcomes of several diseases, but so far such parameters have not been established to detect disease progression in MG.

**Methods:** We conducted a retrospective analysis of selected laboratory parameters related to inflammation and hemogram for MG patients with MC matched to MG patients without MC. To identify potential risk factors for MC, we applied time-varying Cox regression for time to MC, and as a sensitivity analysis generalized estimating equations logistic regression for the occurrence of MC at the next patient visit.

**Results:** 15 of the 58 examined MG patients suffered at least one MC. There was no notable difference in the occurrence of MC by antibody status or sex. Both regression models showed that higher counts of basophils (per 0.01 units increase: HRlJ=lJ1.32, 95% CIlJ=1.02-1.70), neutrophils (per 1 unit increase: HRlJ=lJ1.40, 95% CIlJ=lJ1.14-1.72), and potentially leukocytes (per 1 unit increase: HR = 1.15, 95% CI = 0.99-1.34), and platelets (per 100 units increase: HR = 1.54, 95% CI = 0.99-2.38) may indicate increased risk for a myasthenic crisis.

**Conclusion:** This pilot study provides proof of concept that increased counts of basophils, neutrophils, leukocytes, and platelets may be associated with a higher risk for developing MC in patients with MG.

## INTRODUCTION

Myasthenia gravis (MG) is a rare autoimmune disease caused by an antibody-mediated disturbance of signal transduction at the neuromuscular endplate. The main symptoms are fatigable weakness of the voluntary muscles, worsening with exertion, and fatigue (1). In 70-80% of all patients, MG is caused by pathogenic autoantibodies directed against the acetylcholine receptor (AChR) at the neuromuscular junction (2–4). The loss of functional AChR leads to a reduced amplitude of the endplate potential and thus to impeded neurotransmission at the neuromuscular endplate (5, 6). MG manifests at the extraocular muscles, leading to ptosis or double vision, and by generalized or bulbar weakness affecting limb muscles or oropharyngeal muscles at manifestation or as the disease progresses.

Critical exacerbation of these symptoms can lead to life-threatening myasthenic crisis (MC), which often requires intensive care treatment with non-invasive or even invasive ventilation, and invasive therapy (7). An MC often occurs within the first few years of the disease and can be the first manifestation of MG (8). Lifetime prevalence of MC is 15-20% for patients with MG (9, 10). MC-associated mortality is commonly reported between 5% and 12% (10–13), but mortality up to 22% has also been reported (14, 15). Furthermore, it is established that antibody status or clinical treatment protocols are associated with outcome after MC (10, 13, 16, 17).

Although it is known that certain drugs, inadequate treatment, surgery, infection, sepsis, and pregnancy can trigger MC (9, 18, 19), the prediction of severe exacerbation of MG or ultimately MC based on laboratory parameters is currently not possible. To this end, so far only a few studies investigated the relationship between hemogram or inflammation-related laboratory parameters and disease progression of MG (20–22).

Thus, we hypothesized that certain laboratory parameters could be used to evaluate disease activity in MG even before clinically obvious exacerbation, and to identify patients at risk of progressing to MC. We studied highly granular laboratory parameters related to inflammation and hemogram in patients suffering from MC prior to the event compared to MG without MC to investigate if changes in these parameters could be indicative for development of MC. This retrospective case-control study with a small number of subjects serves as a pilot study, whose concept and results could later be validated in a larger cohort.

## MATERIAL AND METHODS

### Standard protocol approvals, registrations, and patient consent

This study was approved by the ethics committee at Charité – Universitätsmedizin Berlin (no. EA4/068/22). Data were collected retrospectively. Due to the retrospective nature, individual patient consent was not obtained, in accordance with the ethics approval, and state and national laws.

### Study design and patient selection

For this study we evaluated clinical data from 58 MG patients treated at the integrated Myasthenia Center of the Department of Neurology at Charité – Universitätsmedizin Berlin. It is certified by the German Myasthenia Gravis Society and employs standardized workflows for patient management. Diagnosis of MG was established based on antibody studies, repetitive nerve stimulation, or clinical assessment. MC was defined as exacerbation of myasthenic symptoms with bulbar or general weakness requiring mechanical ventilation. First, we selected 15 patients who were treated for MC at least once and for whom sufficiently complete medical data were available from all MC patients at our center between 2006 and 2016. MC patients were intended to be matched in a 1:3 ratio with MG patients treated at our center without recorded MC until 2018. Although data for MC patients were available after 2016, they were not included in the analysis to avoid hind-sight bias. Matching was based on the criteria sex, age ± 5 years, antibody status (AChR antibodies or negative for AChR, MuSK, LRP4), thymectomy (yes/no) and thymus pathology (thymoma, thymus hyperplasia, unremarkable). Due to insufficient matching partners with applicable matching criteria, one MC patient could only be matched with one control patient. The final cohort consisted of 58 subjects – 15 MC, 43 non-MC patients.

Here, we focused on the analysis of the following laboratory parameters: hemoglobin, hematocrit, mean corpuscular hemoglobin (MCH), mean corpuscular hemoglobin concentration (MCHC), white blood cell count, as well as white blood cell differential count (basophils, eosinophils, monocytes, lymphocytes, neutrophils, and granulocytes), platelet count, and C-reactive protein (CRP). Selected laboratory data were obtained through the Berlin Institute of Health at Charité Health Data Platform (HDP), which hosts up-to-date retrospective data of the hospital management system. For this pilot study, we retrieved all available data for the curated list of laboratory parameters of the selected patients over the entire observation period from 2006 to 2018. For the analysis, we only considered data obtained prior to the occurrence of an MC.

### Statistical Analysis

We descriptively display all patient characteristics used for matching, separately for cases and controls. Categorical variables are presented as absolute and relative frequencies. To summarize the laboratory parameters, we display the first measurement per patient (baseline), as well as the median value per patient measured before the beginning of the first MC with median and interquartile range (IQR). Kaplan-Meier curves display the time to first MC stratified by sex (male or female) and antibody status (AChR positive or negative).

We used an Anderson-Gill model, a time-varying Cox proportional-hazards regression, with time to MC as the outcome. To account for the dependency in the data, we used robust standard errors. Time was modelled since first observation, patients without any further crisis were censored at time of database excerpt. This model assumes that the risk to experience an MC remains the same irrespective of whether previous events occurred or not. This means that after an MC has occurred, a subject is treated the same way as a subject who has not experienced an MC. As sensitivity analysis, we also performed a generalized estimating equations (GEE) logistic regression model, which explains the binary outcome of a potential current crisis with the laboratory parameter measured at the prior visit.

Because of the initial matching, we do not adjust for age and sex in any of the models. Due to the limited number of observed events, all models were ran for each laboratory parameter separately (univariable models). Using complete-case analyses, these models are therefore based on a different number of observations, due to the clinical practice not to measure all laboratory parameters at every time point. Based on these models, we derived hazard ratios (HR) and odds ratio (OR) estimates along with 95% confidence intervals (CI). All analyses were performed using R (R Project for Statistical Computing (23)), as well as additional R packages for data handling and analysis (24–26).

## RESULTS

### Demographics and clinical characteristics

This pilot study included 58 patients (30 female, 28 male), of whom 15 (26%) suffered from one or more MC (cases) and 43 never had an MC (controls) within the observation period. In the case group, 11 patients suffered one, two patients two, and two patients three MCs. In total there were 21 MC events (Table 1). Both baseline (i.e., first-ever recorded) and median values for all available measurements prior to MC were similar for all laboratory parameters in both groups. CRP differed in cases and controls (Table 2). The frequency of measurements per person and laboratory parameter varied. For the controls, the median number of observations was 8 (IQR: 3-26, min = 1, max = 714), for cases 42 (IQR: 18-67.5, min = 1, max = 101). The median number of measurements for the complete blood count with differential was 8 (IQR: 2-19.5, min: 1, max: 56), and 15 (IQR: 4-33, min: 1, max: 158) without differential. For CRP it was 6 (IQR: 2-15.5, min: 1, max: 93). The median time between two measurements was 3 days (IQR: 2-35, min: 1, max: 3135). Stratified by the outcome, for the 1944 observations where no MC occurred in the subsequent visit, the median time to next visit (i.e., time to next measurement) was 3 days (IQR: 2-34, min: 1, max: 3135), and 39 days (IQR: 19.2-83.8, min: 1, max: 2196) for the 20 observations where an MC occurred.

**Table 1:** Cohort demographics and clinical characteristics.

|  | **Myasthenic Crisis**<br>**(n=15)** | **No Myasthenic Crisis**<br>**(n=43)** |
| --- | --- | --- |
| **sex** |  |  |
| \| male, n (%) | 7 (46.7) | 21 (48.8) |
| \| female, n (%) | 8 (53.3) | 22 (51.2) |
| **Age** |  |  |
| Age at diagnosis, years, mean (SD) | 56 (16.5) | 51 (17.2) |
| Early onset MG, n (%) | 6 (40) | 16 (37.2) |
| Late onset MG, n (%) | 9 (60) | 27 (62.8) |
| **number of MC per patient** |  |  |
| \| 0 | 0 (0.0%) | 43 (100.0%) |
| \| 1 | 11 (73.3%) | 0 (0.0%) |
| \| 2 | 2 (13.3%) | 0 (0.0%) |
| \| 3 | 2 (13.3%) | 0 (0.0%) |
| **AChR antibodies** |  |  |
| \| positive | 10 (66.7%) * | 28 (65.1%) |
| \| negative for AChR, MuSK, LRP4 | 5 (33.3%) | 15 (34.9%) |
| **thymectomy** |  |  |
| \| no, n (%) | 5 (33.3%) | 15 (34.9%) |
| \| yes, n (%) | 10 (66.7%) | 28 (65.1%) |
| **thymus pathology** |  |  |
| thymoma, n (%) | 5 (50%) | 14 (50%) |
| hyperplasia, n (%) | 2 (20%) | 5 (18%) |
| unremarkable, n (%) | 3 (30%) | 8 (29%) |
| unknown, n (%) | - | 1 (4%) |
| *One MC patient was found positive for AChR and MuSK. As there were no appropriate AChR/MuSK-double positive controls, she was considered in the AChR+ group. |  |  |

**Table 2:** Hemogram and inflammation-related laboratory parameter measurements of patients. For each laboratory parameter, the baseline (first measurement recorded per patient) and the median of all measurements recorded within the observation period before the beginning of the first MC are shown. Missing data are indicated where observed.

|  | MC (n=15) | no MC (n=43) |
| --- | --- | --- |
| **basophils/nl (baseline)** |  |  |
| \| Median (IQR) | 0.03 (0.02, 0.05) | 0.03 (0.02, 0.05) |
| \| Missing | 1 (6.67%) | 1 (2.33%) |
| **basophils/nl (median)** |  |  |
| \| Median (IQR) | 0.03 (0.02, 0.05) | 0.04 (0.02, 0.05) |
| \| Missing | 1 (6.67%) | 1 (2.33%) |
| **C-reactive protein in mg/l (baseline)** |  |  |
| \| Median (IQR) | 7.10 (4.05, 14.93) | 3.50 (1.37, 8.07) |
| \| Missing | - | 19 (44.19%) |
| **C-reactive protein in mg/l (median)** |  |  |
| \| Median (IQR) | 12.00 (4.20, 34.42) | 4.35 (1.68, 8.02) |
| \| Missing | - | 19 (44.19%) |
| **eosinophils/nl (baseline)** |  |  |
| \| Median (IQR) | 0.06 (0.03, 0.10) | 0.08 (0.03, 0.14) |
| \| Missing | 3 (20%) | 3 (6.98%) |
| **eosinophils/nl (median)** |  |  |
| \| Median (IQR) | 0.07 (0.06, 0.12) | 0.08 (0.06, 0.14) |
| \| Missing | 3 (20%) | 3 (6.98%) |
| **hematocrit in l/l (baseline)** |  |  |
| \| Median (IQR) | 0.43 (0.40, 0.46) | 0.42 (0.40, 0.44) |
| **hematocrit in l/l (median)** |  |  |
| \| Median (IQR) | 0.41 (0.37, 0.43) | 0.42 (0.38, 0.43) |
| **hemoglobin in g/dl (baseline)** |  |  |
| \| Median (IQR) | 14.20 (13.55, 15.65) | 14.10 (13.40, 14.65) |
| **hemoglobin in g/dl (median)** |  |  |
| \| Median (IQR) | 13.40 (12.15, 14.55) | 13.80 (12.85, 14.60) |
| **leukocytes/nl (baseline)** |  |  |
| \| Median (IQR) | 9.20 (7.28, 13.19) | 8.20 (6.06, 10.28) |
| \| Missing | 0 (0%) | 0 (0%) |
| **leukocytes/nl (median)** |  |  |
| \| Median (IQR) | 8.11 (6.99, 10.25) | 7.60 (6.65, 8.96) |
| **lymphocytes/nl (baseline)** |  |  |
| \| Median (IQR) | 1.57 (1.08, 1.64) | 1.52 (1.05, 1.88) |
| \| Missing | 1 (6.67%) | 1 (2.33%) |
| **lymphocytes/nl (median)** |  |  |
| \| Median (IQR) | 1.30 (0.88, 1.80) | 1.33 (1.04, 1.71) |
| \| Missing | 1 (6.67%) | 1 (2.33%) |
| **mean corpuscular hemoglobin in pg (baseline)** |  |  |
| \| Median (IQR) | 30.80 (29.05, 32.00) | 30.70 (29.85, 31.75) |
| **mean corpuscular hemoglobin in pg (median)** |  |  |
| \| Median (IQR) | 30.70 (29.45, 31.60) | 30.60 (29.35, 31.80) |
| **mean corpuscular hemoglobin concentration in g/dl (baseline)** |  |  |
| \| Median (IQR) | 33.30 (33.05, 34.35) | 33.90 (33.30, 34.50) |
| **mean corpuscular hemoglobin concentration in g/dl (median)** |  |  |
| \| Median (IQR) | 32.80 (32.60, 33.55) | 33.60 (32.75, 34.33) |
| **monocytes absolute/nl (baseline)** |  |  |
| \| Median (IQR) | 0.59 (0.25, 0.90) | 0.51 (0.42, 0.66) |
| \| Missing | 1 (6.67%) | 1 (2.33%) |
| **monocytes absolute/nl (median)** |  |  |
| \| Median (IQR) | 0.63 (0.47, 0.81) | 0.56 (0.49, 0.69) |
| \| Missing | 1 (6.67%) | 1 (2.33%) |
| **neutrophils/nl (baseline)** |  |  |
| \| Median (IQR) | 6.98 (4.88, 10.75) | 5.43 (3.81, 8.01) |
| \| Missing | 1 (6.67%) | 1 (2.33%) |
| **neutrophils/nl (median)** |  |  |
| \| Median (IQR) | 6.40 (5.41, 7.89) | 5.35 (4.25, 6.31) |
| \| Missing | 1 (6.67%) | 1 (2.33%) |
| **platelets/nl (baseline)** |  |  |
| \| Median (IQR) | 268.00 (220.50, 324.00) | 246.00 (206.50, 283.50) |
| **platelets/nl (median)** |  |  |
| \| Median (IQR) | 260.00 (214.50, 297.00) | 249.50 (210.25, 279.50) |
| **immature granulocytes/nl (baseline)** |  |  |
| \| Median (IQR) | 0.04 (0.02, 0.06) | 0.03 (0.01, 0.04) |
| \| Missing | 1 (6.67%) | 1 (2.33%) |
| **immature granulocytes/nl (median)** |  |  |
| \| Median (IQR) | 0.04 (0.03, 0.07) | 0.03 (0.02, 0.05) |
| \| Missing | 1 (6.67%) | 1 (2.33%) |

### Time to first myasthenic crisis stratified by sex and antibody status

We calculated Kaplan-Meier curves for time to first MC since the first recorded laboratory parameter, stratified by AChR antibody status and sex. Overall, there was neither a significant difference in the occurrence of MC depending on antibody status (Figure 1a) nor between women and men (Figure1b).

**Figure 1:**
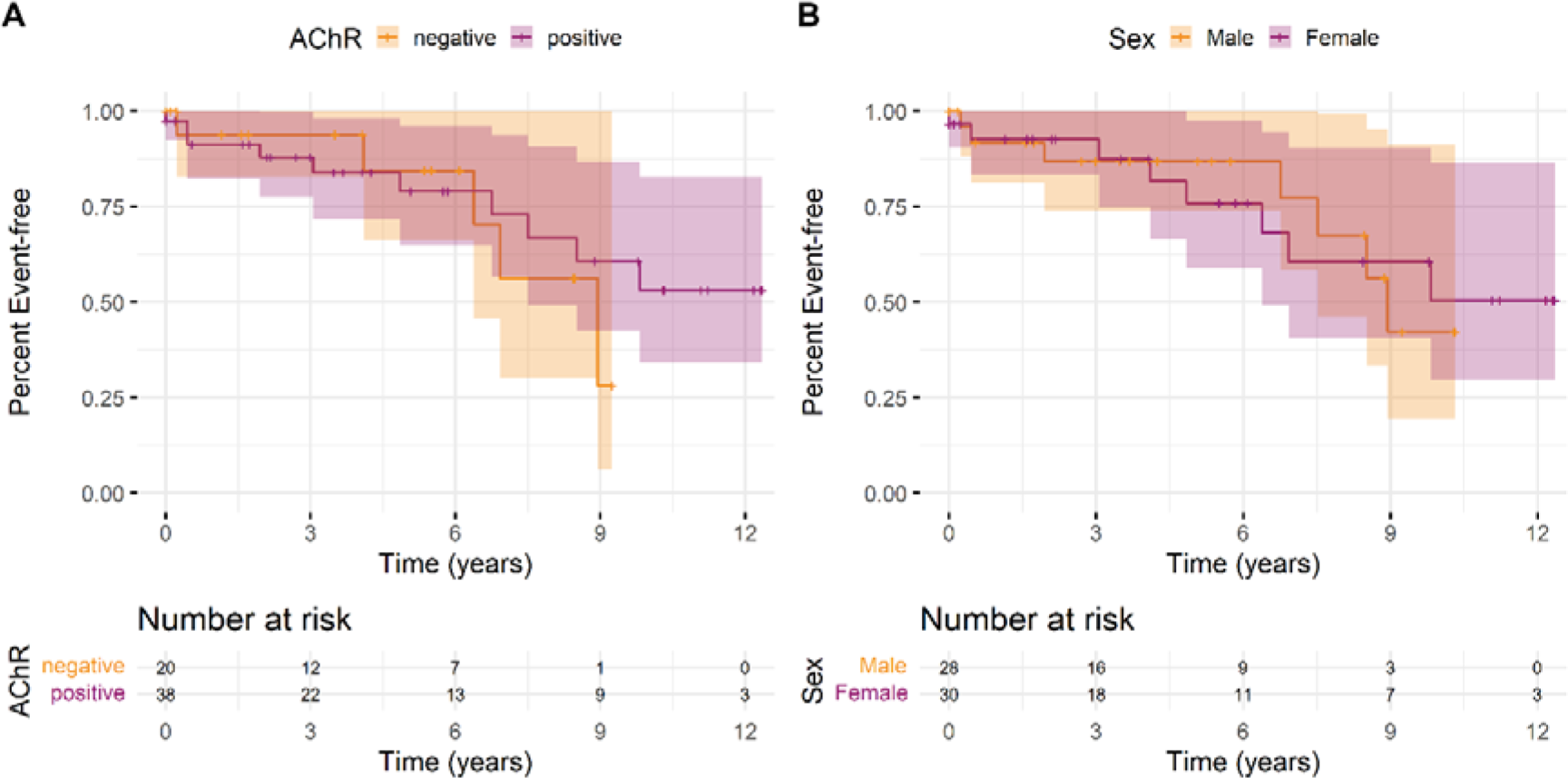
Kaplan Meier curves cohort with 95% confidence intervals (shaded areas) for time to first myasthenic crisis since the first recorded laboratory parameter stratified by (a) antibody status or (b) sex over the observation period. An event is defined as the first MC of a patient. The individual lines show the stratified survival curves, the shaded areas present the 95% confidence interval. (a) Orange lines represent the data for male and purple for female patients. (b) Orange lines show the data for AChR antibody negative patients, and purple lines for AChR antibody positive patients. Vertical bars indicate censoring of an observation at that point. Additionally, the number of individuals at risk for a first MC at regular time points is shown below the curves.

### Laboratory parameters associated with myasthenic crisis

Both statistical models we applied make use of the previous measurement to explain the occurrence of an event (MC or no MC). Laboratory parameter measurements from 1964 observations were used to explain 20 events with subsequent MC (1 MC did not have sufficiently complete data to be included), and 1944 observations without subsequent MC. Univariable Anderson-Gill models showed that basophils, neutrophils, and potentially leukocytes and platelets indicate increase hazards for a myasthenic crisis (Figure 2). Without adjustment for other parameters, an increase of basophils by 0.01 units increased the risk of an MC 1.32-fold (95% CI: 1.02-1.70), a 1-unit increase in neutrophils 1.4-fold (95% CI: 1.14-1.72). Furthermore, every unit increase in leukocytes increased the hazard for MC 1.15-fold (95% CI: 0.99-1.34), an increase of 100 units in platelets 1.54-fold (95% CI: 0.99-2.38).

**Figure 2:**
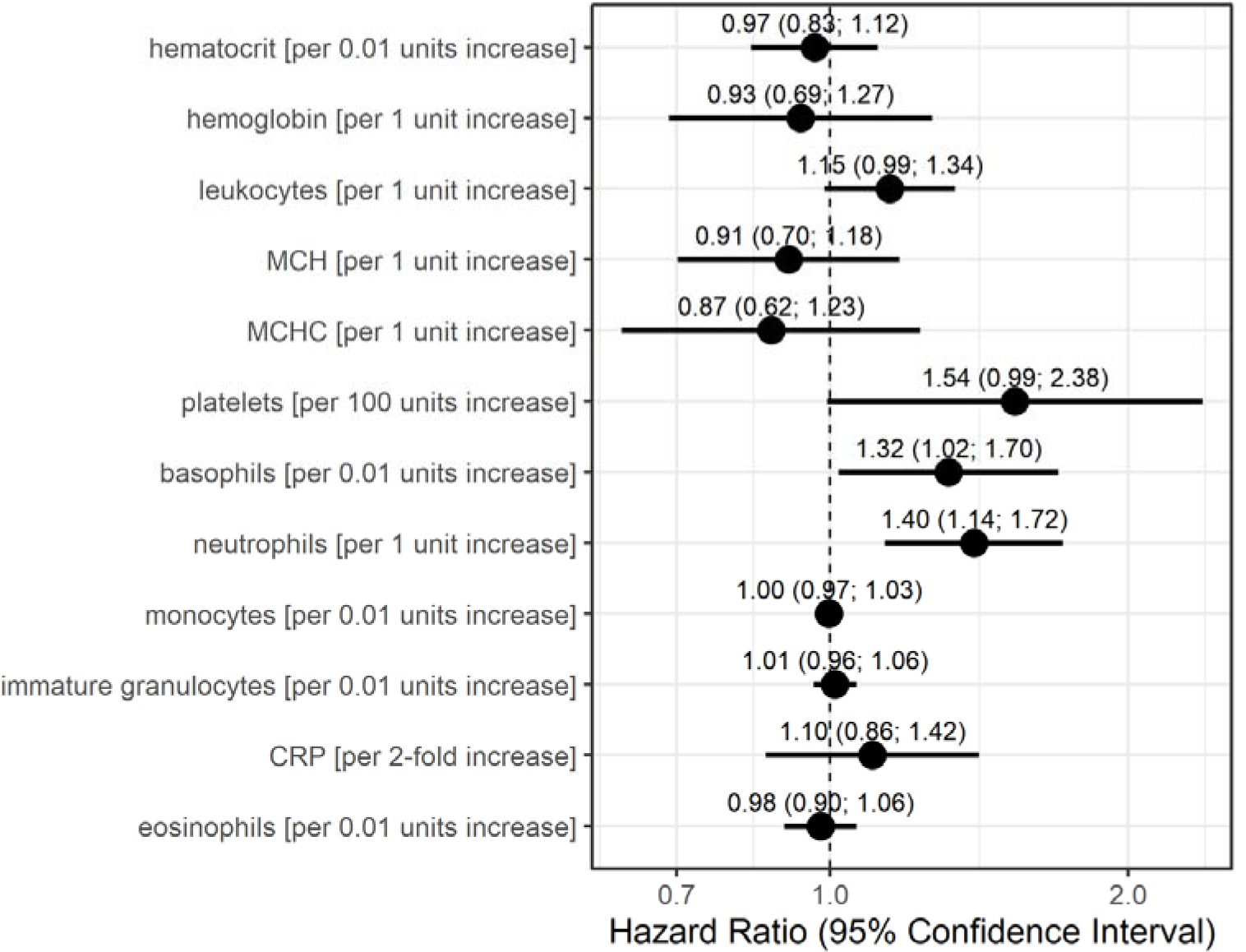
Hazard Ratio estimates with 95% confidence intervals based on Anderson-Gill models for each laboratory parameter, for occurrence of myasthenic crisis. Dots indicate hazard ratio estimates for each laboratory parameter, and horizontal bars 95% confidence intervals (in parentheses). Units for each laboratory parameter are shown in Table 2.

The GEE logistic regression models conducted as sensitivity analyses with occurrence of an MC in the subsequent patient visit as the outcome also identified basophils, neutrophils, and platelets as potentially relevant laboratory parameters (Figure 3). The odds for MC in the subsequent visit were 1.27-fold (95% CI:1.08-1.49) per 0.01 unit increase in basophils, and 1.15-fold (95% CI: 1.02-1.30) per 1 unit increase in neutrophils. A 100 units increase in platelets increased the odds for an event 1.29-fold (95% CI: 0.85-1.95), although this association was calculated with low precision. Additionally, higher values in hematocrit (per 0.01 units) and hemoglobin (per 1 unit) resulted in higher odds for a subsequent MC (OR = 1.11, 95% CI: 1.0.1-1.22, OR = 1.19, 95% CI: 1.01-1.39, respectively).

**Figure 3:**
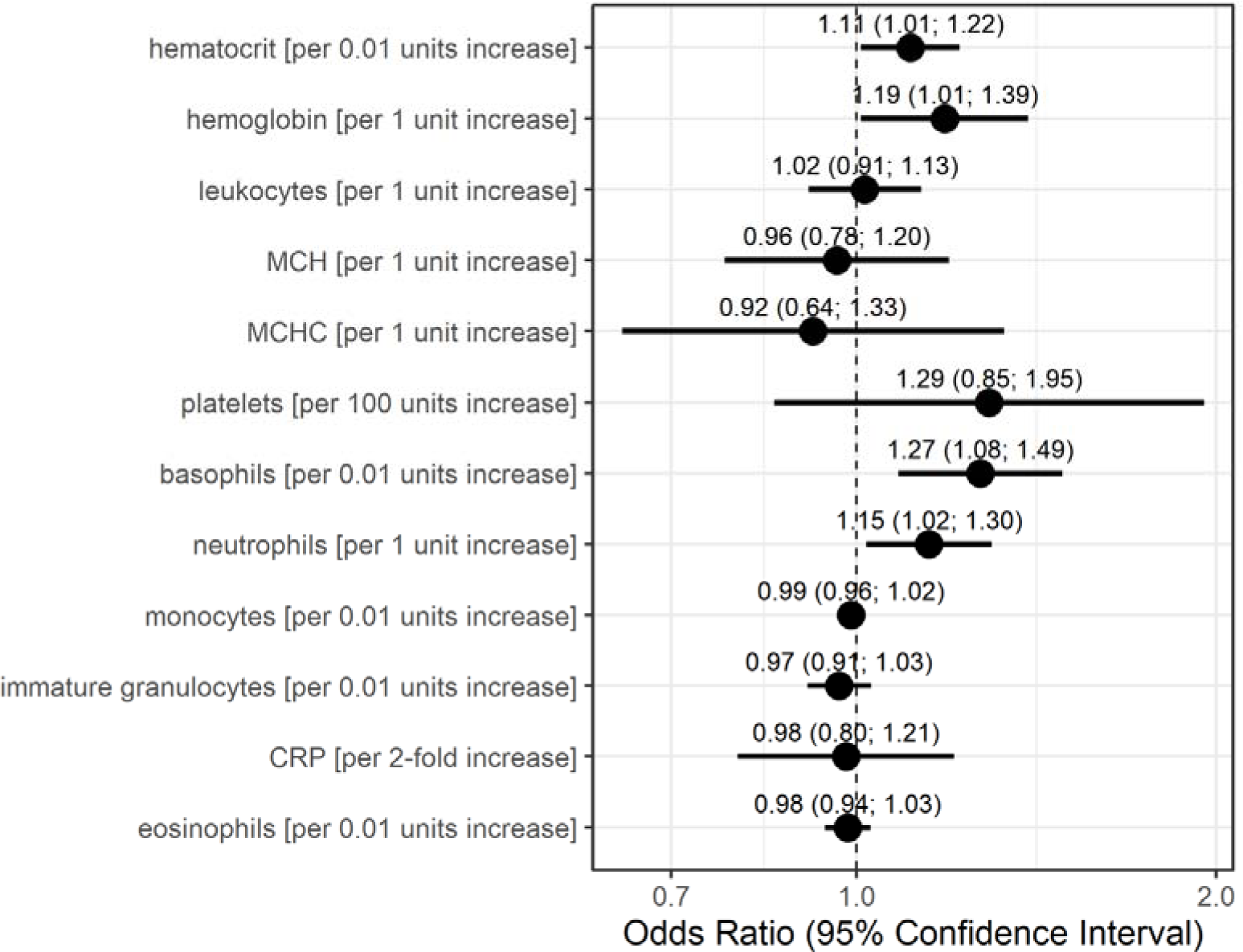
Odds Ratio estimates and 95% confidence intervals based on generalized estimating equations logistic regression for each individual laboratory parameter, for occurrence of myasthenic crisis at the subsequent visit. Dots indicate odds ratio estimates for each laboratory parameter, and horizontal bars indicate 95% confidence intervals (in parentheses). Estimates were derived from the generalized estimating equations (GEE) logistic regression. Units for each laboratory parameter are shown in Table 2.

Both statistical models consider data only before the occurrence of an MC but differently account for time. The Anderson-Gill model is a time-varying Cox regression model and as such explores the relationship between the time to the occurrence of an event and the explanatory variables. The dependent variable here is the hazard function at a given time t. Therefore, the model is dependent on time, as the hazard of an MC to occur changes with time. The GEE logistic regression estimates the odds for an event (here the occurrence of an MC at the next visit) based on the explanatory variables in the model. Time is only taken into account by the sequence of measurements and the occurrence of an MC at the next visit. Based on the results of these models we conclude that increased values of basophils, neutrophils, and with lower confidence also leukocytes and platelets are associated with an increased risk of MC.

## DISCUSSION

In this pilot study, we investigated whether routine laboratory data could be used to anticipate occurrence of MC during the disease course in MG patients. Based on two statistical models with distinct assumptions we found that from our pre-selected set of laboratory parameters higher basophils, neutrophils, leukocytes, and platelets measured before an event (i.e., MC) were associated with a higher risk of developing an MC. Although baseline and median CRP before MC was elevated in the MC group compared to the control group, it was not identified with a risk to develop an MC by either statistical model.

Several risk factors for MC have previously been identified from retrospective analyses: infections (15, 19, 27), drugs (27), corticosteroid treatment (28, 29), older age (30), thymoma (15), bulbar symptoms (15, 31), high disease severity (15, 30), male sex (15, 29), and the presence of additional autoimmune diseases (30, 31). Another risk factor for MC is surgery (15, 32), including thymectomy. Although some attempts to establish risk scores for the occurrence of postoperative myasthenic crisis have been made, the identified risk factors (bulbar symptoms, disease severity, decreased vital capacity, thymoma) generally align with established general risk factors for MC (32–34). Due to the high mortality rate of MC of 5-12% (10, 13, 16, 18), which can be stratified by AChR (10) or MuSK (16) antibodies, and triple-seronegative patients (13), there is a significant need to establish risk scores or to identify parameters which can be used to predict the occurrence of MC, and thus aid early intervention.

Our study provides initial indications that routine laboratory parameters assessed before the onset of MC could be used as risk predictors for MC occurrence and facilitate early interventions (e.g., treatment with immunoglobulins or plasma exchange), possibly preventing MC and mitigate the associated morbidity and mortality. To this end, some studies have investigated prediction of in-hospital mortality in MC based on selected laboratory parameters (22, 35). A recent study derived a predictive score for in-hospital mortality of MC which used Myasthenia Gravis Foundation of America (MGFA) score at onset of the MC, septic shock, and cardiac arrest (35). In addition, this study suggested that low serum albumin, low hemoglobin, and high leukocyte count might be associated with a higher mortality in MC (35). The latter may corroborate our findings that increased leukocyte counts may be associated with increased risk for an ensuing MC. Further studies described a possible association between infections (36) and signs of inflammation (leukocytosis) (22) with an increased risk of developing MC. Furthermore, hemogram could provide clues to the course of the disease, as hematological changes have been identified as prognostic factors of mortality for several critical illnesses (22), e.g., endocarditis (37), acute kidney injury (38), and acute myocardial infarction (39, 40). Extreme leukocytosis and anemia have been described as important risk factors for increased mortality in MC (22). Furthermore, elevated neutrophil-to-lymphocyte ratios have been reported to be a potential risk factor for indicating disease severity of MG in children (20) and in adults (21). Furthermore, a recent study used explainable machine learning to classify the risk for MC based on real world clinical data, including laboratory results (41). Thus, to estimate the risk of developing MC in a given patient, routine laboratory parameters could be used as they are inexpensive and widely available.

There are several limitations to our study. The dataset with 58 patients in total is small. However, the dataset includes 21 MC events and several sequential laboratory measurements per patient since the laboratory parameters were measured frequently. This leads to an uneven distribution of measurements between cases and controls. Furthermore, there is the potential of selection bias due to the retrospective and mono-centric design and hand-selection of cases and controls for this pilot study. This study did not consider further clinical data such as information on infection or co-medication, which are known risk factors for clinical worsening of MG. Likewise, steroids or steroid-sparing immunosuppression are standard medications in MG patients known to affect blood counts but were not considered as confounders. We used complete-case analyses based on a different number of observations, as a result of clinical practice not to measure all laboratory parameters at every time point. This leads to a different number of measurements per parameter per patient, and thus they could only be considered as univariate parameters in the models.

In conclusion, this study indicates that increased basophils, neutrophils, leukocytes, and platelets may be associated with an increased risk for the occurrence of MC in MG patients. The results of this pilot study suggest that it is possible to identify predictors for MC risk based on routine laboratory data. Together with other medical data (41), routine blood biomarkers could serve to develop a risk prediction score to tailor individualized treatment decisions at the point of care. However, larger prospective studies beyond the proof-of-concept stage are necessary to verify our results.

## Data availability statement

Ethical approval currently does not permit sharing of raw data. Approval will be sought by the corresponding author upon reasonable request with scientific rationale and sound methodology. Requests for data sharing will be managed in accordance with data access and sharing policies of Charité – Universitätsmedizin Berlin.

## Author contributions

A. Mehnert collected, curated, discussed and interpreted the data, wrote the initial draft and edited manuscript. S.B. performed data analyses, and discussed and interpreted the data. J.K.-H. performed statistical analyses, and discussed and interpreted the data. L.G., M.L., S.H., S.L., F.Sch., F.St., M.S., A.M.B. discussed the data and edited the manuscript for intellectual content. A. Meisel discussed and interpreted the data, and edited the manuscript for intellectual content. A.A. devised, performed, and supervised the statistical analyses, discussed and interpreted the data, and edited the paper. P.M. conceived and supervised all aspects of the study, discussed and interpreted the data, and revised and edited the manuscript. All authors contributed to the submitted version of the paper and approved it for publication.

## Acknowledgements

We thank C. Heibutzki, D. Remstedt, J. Brestrich and N. Baro for study and patient management, S. Märschenz, S. Lischewski, and M. Heinold for administrative support, and M. Mandrela, P. Brunecker and the team of the Health Data Platform at the Berlin Institute of Health at Charité for data access support.

